# Latin American Trans-ancestry INitiative for OCD genomics (LATINO): Study Protocol

**DOI:** 10.1101/2023.02.23.23286373

**Authors:** James J Crowley, Carolina Cappi, Marcos E Ochoa-Panaifo, Renee M Frederick, Minjee Kook, Andrew D Wiese, Diana Rancourt, Elizabeth G Atkinson, Paola Giusti-Rodriguez, Jacey L Anderberg, Latin American Trans-ancestry INitiative for OCD genomics (LATINO), Brazilian Obsessive-Compulsive Spectrum Disorder Working Group (GTTOC), Jonathan S Abramowitz, Victor R Adorno, Cinthia Aguirre, Gustavo S Alves, Gilberto S Alves, NaEshia Ancalade, Alejandro A Arellano Espinosa, Paul D Arnold, Daphne M Ayton, Izabela G Barbosa, Laura Marcela Barón Castano, Cynthia N Barrera, María Belén Prieto, María Celeste Berardo, Dayan Berrones, John R Best, Tim B Bigdeli, Christie L Burton, Jennifer L Callahan, Maria Cecília B Carneiro, Sandra L Cepeda, Evelyn Chazelle, Jessica M Chire, Macarena Churruca Munoz, Pamela Claisse Quiroz, Journa Cobite, Jonathan S Comer, Daniel L Costa, Jennifer Crosbie, Victor O Cruz, Guillermo Dager, Luisa F Daza, Anabel de la Rosa-Gómez, Daniela del Río, Fernanda Z Delage, Carolina B Dreher, Lucila Fay, Tomas Fazio, Ygor A Ferrão, Gabriela M Ferreira, Edith G Figueroa, Leonardo F Fontenelle, Diego A Forero, Daniele TH Fragoso, Bharathi S Gadad, Sheldon R Garrison, Andres González, Laura D Gonzalez, Marco A González, Polaris Gonzalez-Barrios, Wayne Goodman, Jerry Guintivano, Daniel G Guttfreund, Andrew G Guzick, Matthew W Halvorsen, Joseph D Hovey, Reinhard Janssen-Aguilar, Matias Jensen, Alexandra Z Jimenez Reynolds, Joali Alexandra Juárez Lujambio, Nasim Khalfe, Madison A Knutsen, Caleb Lack, Nuria Lanzagorta, Monicke O Lima, Melanie O Longhurst, David A Lozada Martinez, Elba S Luna, Andrea H Marques, Molly Martinez, Maria de Los Angeles Matos, Caitlyn E Maye, Joseph F McGuire, Gabriela Menezes, Charlene Minaya, Tomás Miño, Sara M Mithani, Circe Montes de Oca, Alonso Morales-Rivero, Maria E Moreira-de-Oliveira, Olivia J Morris, Sandra I Muñoz, Zainab Naqqash, Ambar A Núñez Bracho, Belinda E Núñez Bracho, Maria Corina Ochoa Rojas, Luis A Olavarria Castaman, Iliana Ortega, Darpan I Patel, Ainsley K Patrick, Mariel Paz y Mino, Jose L Perales Orellana, Bárbara Perdigão Stumpf, Tamara Peregrina, Tania Pérez Duarte, Kelly L Piacsek, Maritza Placencia, Lucas C Quarantini, Yana Quarantini-Alvim, Renato T Ramos, Iaroslava C Ramos, Vanessa R Ramos, Kesley A Ramsey, Elise V Ray, Margaret A Richter, Bradley C Riemann, Juan C Rivas, Maria C Rosario, Camilo J Ruggero, Angel A Ruiz-Chow, Alejandra Ruiz-Velasco, Aline S Sampaio, Leonardo C Saraiva, Russell J Schachar, Sophie C Schneider, Ethan J Schweissing, Laura D Seligman, Roseli G Shavitt, Keaton J Soileau, S. Evelyn Stewart, Shaina B Storch, Emily R Strouphauer, Kiara R Timpano, Beatriz Treviño-de la Garza, Javier Vargas-Medrano, María I Vásquez, Guadalupe Vidal Martinez, Saira A Weinzimmer, Mauricio A Yanez, Gwyneth Zai, Lina M Zapata-Restrepo, Luz M Zappa, Raquel M Zepeda-Burgos, Anthony W Zoghbi, Euripedes C Miguel, Carolyn I Rodriguez, Mayra C Martinez Mallen, Pablo R Moya, Tania Borda, María Beatriz Moyano, Manuel Mattheisen, Stacey Pereira, Gabriel Lázaro-Muñoz, Karen G Martinez-Gonzalez, Michele T Pato, Humberto Nicolini, Eric A Storch

## Abstract

Obsessive-compulsive disorder (OCD) is a debilitating psychiatric disorder. Worldwide, its prevalence is ~2% and its etiology is mostly unknown. Identifying biological factors contributing to OCD will elucidate underlying mechanisms and might contribute to improved treatment outcomes. Genomic studies of OCD are beginning to reveal long-sought risk loci, but >95% of the cases currently in analysis are of homogenous European ancestry. If not addressed, this Eurocentric bias will result in OCD genomic findings being more accurate for individuals of European ancestry than other ancestries, thereby contributing to health disparities in potential future applications of genomics. In this study protocol paper, we describe the Latin American Trans-ancestry INitiative for OCD genomics (LATINO, www.latinostudy.org). LATINO is a new network of investigators from across Latin America, the United States, and Canada who have begun to collect DNA and clinical data from 5,000 richly-phenotyped OCD cases of Latin American ancestry in a culturally sensitive and ethical manner. In this project, we will utilize trans-ancestry genomic analyses to accelerate the identification of OCD risk loci, fine-map putative causal variants, and improve the performance of polygenic risk scores in diverse populations. We will also capitalize on rich clinical data to examine the genetics of treatment response, biologically plausible OCD subtypes, and symptom dimensions. Additionally, LATINO will help elucidate the diversity of the clinical presentations of OCD across cultures through various trainings developed and offered in collaboration with Latin American investigators. We believe this study will advance the important goal of global mental health discovery and equity.

## 1 RATIONALE

### 1.1 Overview

Genomics is now identifying long-elusive risk genes, distinguishing related disorders, and beginning to guide therapy. Unfortunately, most of these advances primarily benefit individuals of European ancestry (EA), potentially exacerbating existing health disparities (Martin et al., 2019). Furthermore, admixed populations, such as “Latinx” (i.e., Latino and Latina) populations, constitute more than a third of the U.S. population and yet are markedly understudied in medical and psychiatric genetics (Peterson et al., 2019). The reasons for the current Eurocentric bias in genomics include socio-cultural factors (Sanderson et al., 2013), such as more funding opportunities in higher-income countries, and analytical reasons where non-EA samples are often simply excluded from EA-focused analyses. Correcting this disparity will expand our understanding of disease risk from common variation observed among individuals of all ancestries, with particular benefit to understudied Latinx individuals. For example, novel loci are bound to be discovered owing to alleles specific to Latinx populations, which will expand our view of disease pathophysiology. Existing loci can be fine-mapped by taking advantage of linkage disequilibrium differences across ancestries, bringing us closer to identifying causal variants. In addition, polygenic risk scores (PRS: a way of measuring common genetic risk factors) will be more accurate in non-EA individuals if the discovery sample includes their ancestral background. These advances should reduce existing health disparities in genomics.

Obsessive-compulsive disorder (OCD) is a disorder that needs to both markedly diversify its genomic sample and to increase its sample size overall. In the past year, the OCD genome-wide association study (GWAS) sample size has grown from just 2,688 cases to ~50,000 cases (~10,000 from clinics, ~10,000 from biobanks, and ~30,000 self-reported OCD cases from 23andMe), according to the latest, unpublished freeze from the Psychiatric Genomics Consortium (PGC) OCD Working Group (presented in October 2022 at the World Congress of Psychiatric Genetics in Florence, Italy). Unfortunately, the sample is far from diverse – over 95% of the cases are of EA. Preliminary GWAS results, based on EA cases and controls, have yielded the first 30 genome-wide significant hits for OCD and brought OCD GWAS to the point where many more loci will be discovered as sample size increases. One of the next steps will be to perform trans-ancestry meta-analyses; but with just ~2,000 non-EA cases from all over the world, power for these analyses is low. There is a need to collect a large number of non-EA participants and to work together with PGC OCD to perform the most highly powered analysis possible to reduce health disparities and identify both common and ancestry-specific loci.

Including people of Latinx ancestry from areas in North and South America provides a fertile ground for such a collection. The genetic makeup of Latinx individuals has been shaped by a history of extensive admixture between Africans, Europeans, and indigenous peoples (Norris et al., 2018). Thus, the region is ideal to examine the interplay of genetic factors from these three major ancestral groups. Therefore, we assembled a network of over 50 sites in Latin America, the United States, and Canada that treat a large number of ancestrally diverse OCD patients (see ***Figure 1***). We have come together with experts in the clinical aspects of OCD, OCD genetics and the genetics of admixed populations for a new NIH-funded study called the **<u>L</u>**atin **<u>A</u>**merican **<u>T</u>**rans-ancestry **<u>IN</u>**itiative for **<u>O</u>**CD genomics (LATINO, www.latinostudy.org).

**Figure 1.**
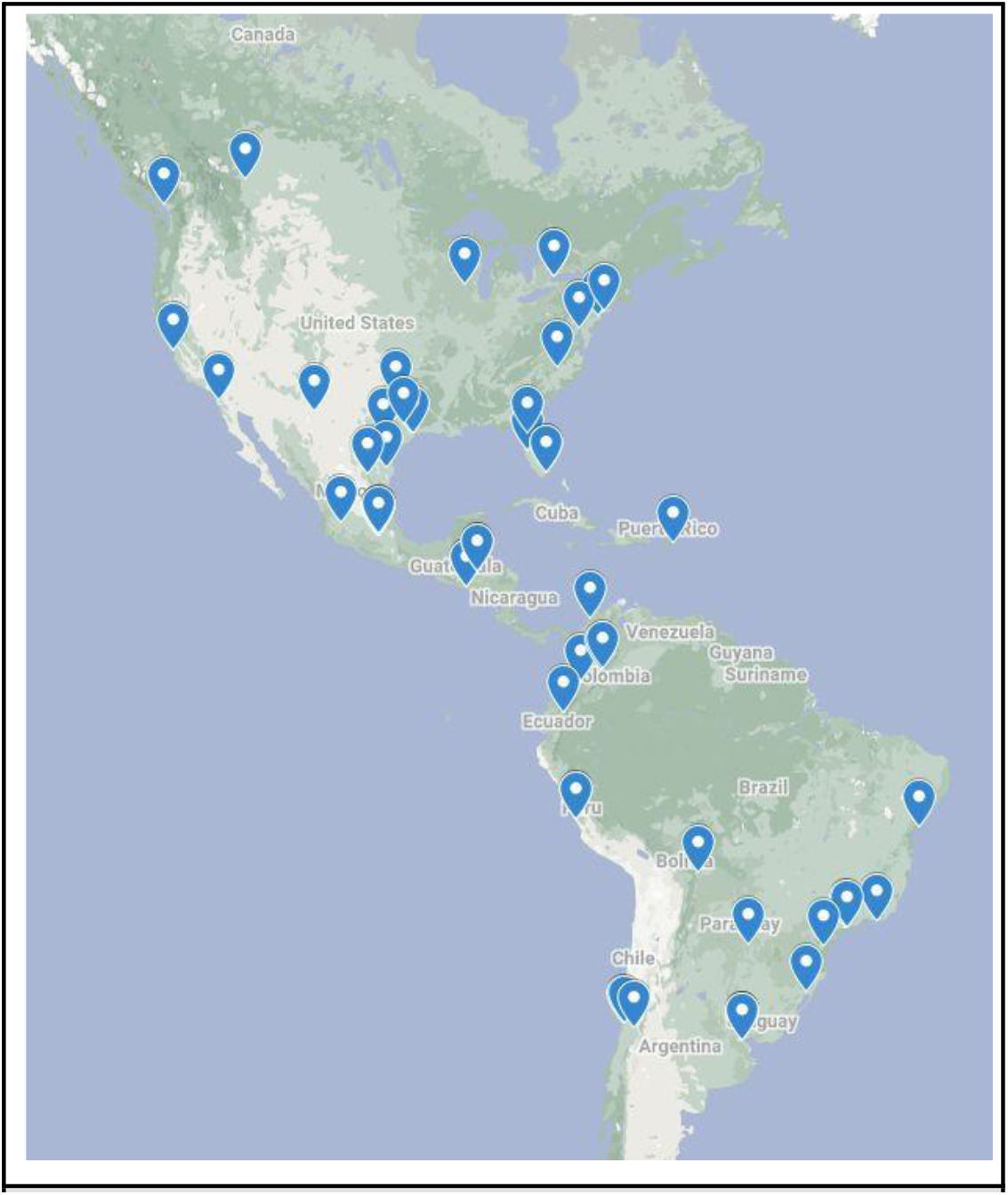
LATINO study sites.

First, LATINO will collect DNA and comprehensive clinical data from at least 5,000 Latinx OCD cases. These cases will be matched with controls collected by other Latinx psychiatric genetics projects. Second, the combined cohort of cases and controls will be subject to genome-wide single nucleotide polymorphism (SNP) genotyping, allowing us to perform a novel trans-ancestry GWAS in collaboration with the PGC OCD Working Group. Third, we will leverage these diverse study data to fine-map new OCD risk loci and compare the performance of the newly diverse PRS with EA-only PRS. We believe this study will highlight the importance of including all major ancestral groups in genomic studies, both as a strategy to improve power to find disease associations, as well as to reduce health disparities, in potential future applications of genomics in precision medicine.

LATINO, however, is much more than a genetic study. OCD is under-recognized, underdiagnosed, under-treated, and under-studied in Latinx individuals, contributing to health disparities (Abramowitz et al., 2017; Benuto, 2017; Perez et al., 2022; Wetterneck et al., 2012). In addition to decreasing the Eurocentric bias in OCD genetics, LATINO is working with stakeholders and patient-centered organizations to increase awareness of OCD in Latinx individuals and improve their access to evidence-based treatments. For example, LATINO is playing a central role in executing social media-based awareness campaigns and providing hands-on training for psychologists and psychiatrists interested in treating OCD. LATINO is also facilitating OCD research in Latin America by translating key instruments into Spanish and Portuguese, validating them in Latinx individuals, and providing a genomics training program. Finally, LATINO is helping Latin American OCD researchers identify funding opportunities and establish mutually beneficial collaborations. To that end, LATINO will actively contribute to decreasing disparities on multiple levels.

### 1.2 Background

OCD is a neuropsychiatric disorder characterized by recurrent, unwanted thoughts, ideas, images or urges and/or repetitive behaviors that are frequently performed in response to the distress associated with the obsessions (American Psychiatric Association, 2013; World Health Organization, 1992). Individuals with OCD experience exaggerated concerns about danger, hygiene or harm that result in persistent conscious attention to the perceived threat (obsessions)(Goodman et al., 2021). In response to distress associated with obsessions, the individual performs compulsions to neutralize the anxiety/distress, which provides temporary relief. However, relief is negatively reinforcing, leading to repetitive, compulsive behavior when obsessions inevitably recur (Pauls et al., 2014). OCD is a multidimensional disorder shown to consist of roughly four primary symptom dimensions (contamination, symmetry, forbidden thoughts and related compulsions, checking) (Bloch et al., 2008) which may have distinct neural circuitry (Mataix-Cols et al., 2004), genetic (Hasler et al., 2007), and etiological origins (Leckman et al., 2009). Additionally, some subtypes such as early-onset and tic-related OCD may be etiologically meaningful subtypes of the disorder (Brander et al., 2021; Leckman et al., 2010).

The OCD lifetime prevalence is ~2% worldwide (Karno et al., 1988; Ruscio et al., 2010; Wetterneck et al., 2012), with onset often in childhood and similar prevalence by sex. Studies of OCD in Latin America have shown similar prevalences (Wetterneck et al., 2012). Most OCD cases have a comorbid psychiatric disorder (e.g., tic, mood, and anxiety disorders) (Fullana et al., 2009). Patients with OCD are at substantial risk of suicide (~10 times higher than the population prevalence)(Fernández de la Cruz et al., 2017). OCD was ranked among the 10 leading causes of years lost to disability in 2008 (Mathers & World Health Organization, 2008). Although medication and behavioral therapy are useful (McGuire et al., 2015), symptom control is imperfect, and the course is often chronic.

A deeper understanding of the pathophysiology of OCD is critical to the development of more effective treatments. Successful completion of our aims will make important contributions to understanding the etiology of this disorder, alongside diversifying the sample to include understudied populations. We believe that the identification of specific genes and biological pathways mediating susceptibility to OCD will lead to improved detection, differential diagnosis, and treatment strategies (Grassi et al., 2020). Our sampling scheme is not only cost-effective; but also captures representative, comprehensively phenotyped Latinx OCD cases in multiple Latin American nations. The participation of trained bioethicists will help guide the collection of samples in a culturally-sensitive and responsible manner.

### 1.3 OCD Genetics

OCD is ~50% heritable (Mataix-Cols et al., 2013; Pauls, 2010; van Grootheest et al., 2005). First-degree relatives of affected individuals have a 4-8x increased risk of OCD (Browne et al., 2015; Mataix-Cols et al., 2013). OCD linkage studies (Pauls et al., 2014), and >100 candidate gene studies (Taylor, 2013), have produced inconsistent results. Regarding rare variation, two key whole exome sequencing (WES) studies have been published. First, Cappi et al (Cappi et al., 2020) performed WES on 222 sporadic OCD cases and their unaffected parents to identify rare *de novo* mutations (DNMs). Results showed that DNMs predicted to damage gene function are enriched in OCD probands. Second, Halvorsen et al (Halvorsen et al., 2021) reported WES results from the largest OCD cohort to date (1,313 total cases, consisting of 587 trios, 41 quartets and 644 singletons). They identified an excess of inherited and *de novo* loss of function (LoF) variation in LoF-intolerant genes and the most significant single-gene result was *SLITRK5* (P = 2.3 × 10^−6^). Taken together, these data support the contribution of rare coding variants to OCD genetic risk.

Regarding common variation, the first well-powered OCD GWAS from the PGC OCD Working Group was posted to medRxiv in October 2021 (Strom et al., 2021). This study was limited to EA individuals (N = 14,140 OCD cases; N = 562,117 controls), identified the first genome-wide significant locus for OCD, and strengthened previous literature suggesting a polygenic nature of this disorder. The PGC OCD Working Group is currently working on publishing a much larger OCD GWAS (~50,000 cases; ~1M controls), but the sample remains heavily European because no large-scale collection of non-European OCD cases has been performed. We expect that LATINO and similar studies of OCD in individuals from diverse ancestries will help to accelerate gene discovery for this understudied condition.

## 2 AIMS AND METHODS

LATINO’s main aims are: 1. Diversify the OCD genomics sample; 2. Discover genomic loci for OCD; and 3. Fine-map OCD loci and test accuracy for PRS (***Table 1***). We describe in detail below the methods proposed to achieve each of these aims.

**Table 1.** LATINO study specific aims

| **Table 1** LATINO study specific aims |
| --- |
| **Aim 1. Diversify OCD genomics sample** |
| A. Collect saliva-derived DNA from 5,000 Latinx OCD cases from a network of clinics across Latin America, the United States and Canada. Matched controls are available through collaboration.<br>B. Perform a detailed phenotypic assessment of each case (e.g., comorbidities, symptom dimensions, severity, treatment response). |
| **Aim 2. Discover genomic loci for OCD** |
| A. Genotype all 5,000 Latinx OCD cases with the Illumina Global Screening Array (GSA) and match with data from at least 10,000 Latinx controls.<br>B. Perform a GWAS on the 5,000 OCD cases and at least 10,000 matched controls and meta-analyze with results from the PGC OCD Working Group. |
| **Aim 3. Fine-map OCD loci and test accuracy of PRS** |
| A. Evaluate fine-mapping gains in terms of reductions in the number of credible SNPs at genome-wide significant loci following trans-ancestry GWAS meta-analysis.<br>B. Compare the performance of PRS trained on our diverse sample with analogous scores based on EA-only samples. |

### 2.1 Aim 1

#### 2.1.1 Cases

Cases must have an ICD-10 diagnosis of OCD (established with the MINI/MINI-KID or SCID), have at least one biological grandparent born in Latin America (Mexico, a Caribbean Island, Central America, or South America) and be between 7 and 89 years of age. A lower limit of 7 years was chosen to allow adequate phenotyping with a range of valid instruments, while an upper limit of 89 years was chosen because anything above this age is considered protected health information in the United States. Case participants can live anywhere in Latin America, the United States, or Canada, and are consented/assessed by investigators in their respective country. All participants are included in the study regardless of psychiatric comorbidity, as long as they fulfill strict diagnostic criteria for OCD. All comorbidity is registered. Patients are excluded in cases of diagnostic uncertainty, such as OCD secondary to a neurological disorder or definite CNS insult, or where the differential diagnosis between OCD and an alternative condition is unclear. Cases are reviewed by site PIs, and a standing caseness panel is held weekly via videoconference to provide a forum to discuss diagnostic issues and questions.

The LATINO network has >50 case recruitment sites at clinics, universities, psychiatric hospitals, day treatment programs, and/or partial hospitalization programs in Latin America, the United States, and Canada (see ***Figure 1***). Participants are being recruited in person at these sites or, in some cases, through a country-specific online screening process (www.latinostudy.org). Web and social media-based advertisements are facilitating recruitment of individuals with OCD who are not engaged in clinical services.

#### 2.1.2 Controls

Controls are inherited from other ongoing psychiatric genetics studies being conducted across Latin America, the United States, and Canada. These studies, like LATINO, are part of the NIMH-funded Ancestral Populations Network (APN). The APN comprises several projects focused on psychiatric genetics in diverse populations and provides LATINO with a resource for ancestry-matched controls screened for psychiatric disorders.

The APN includes multiple relevant studies, such as the Populations Underrepresented in Mental Illness Association Studies (PUMAS)(Lopez-Jaramillo et al., 2022), the Latino Ancestry Genomic Psychiatry Cohort (LA-GPC)(Bigdeli et al., 2020; Pato et al., 2013) and the Neuropsychiatric Genetics in Mexican Populations (NeuroMex) study (Camarena et al., 2021). At least 20,000 controls with Latin American ancestry will be available for matching.

Controls are administered a screening questionnaire composed of 32 questions from well-validated interviews and screening measures for OCD, mania, psychosis, depression, anxiety disorders, and history of alcohol, nicotine and other substance use disorders (Pato et al., 2013). In addition, there is a section specific to demographic information (i.e., age, sex and self-identified race/ethnicity), and a section specific to medical conditions and disorders, including head trauma and seizure history. Individuals reporting no lifetime symptoms indicative of OCD will be considered eligible to be matched control participants.

#### 2.1.3 Approvals and informed consent

Before recruiting any participants, all sites are approved to conduct this study by both the United States Department of State as well as their local Institutional Review Board (IRB). Further, all adult participants will provide informed consent in writing or by using an electronic consenting process approved by their local IRB. All minors will provide their written/electronic consent or verbal assent (depending on their age and local IRB requirements) and written/electronic consent from their parents or legal guardians. When permitted by the local IRB, participants are compensated up to $25 USD after completing the core assessment battery and providing a saliva sample. All participants give permission for their de-identified data to be made available to the scientific community via having the genomic and phenotype data added to repositories (e.g., the U.S. NIMH Data Archive) and shared with international consortia (e.g., PGC). When permitted by the local IRB, an aliquot of DNA will be sent to the NIMH Repository and Genomics Resource (NRGR).

#### 2.1.4 Saliva collection and DNA

All participants donate 2 ml of saliva for DNA. Saliva is collected into SDNA-1000 kits from Spectrum Solutions™ (Draper, Utah, USA), which stabilize the DNA for shipping and storage at room temperature. Participants are instructed not to eat, drink, smoke, or chew gum for 30 minutes prior to collecting the sample. Participants recruited in person are provided a saliva kit for sampling during their appointment, while those recruited online are mailed a saliva kit with a prepaid return envelope.

DNA from 1 ml of saliva is extracted in an automated fashion using Maxwell® RSC Stabilized Saliva DNA Kits and a Maxwell® RSC 48 Instrument from Promega (Madison, Wisconsin, USA). The remaining saliva solution is frozen at -80 degrees C.

#### 2.1.5 Clinical assessments

A strength of this study is the depth and consistency of phenotyping and treatment history, which will yield a genetic study powered to analyze comorbidity, symptom dimensions, and treatment response. All LATINO sites have harmonized diagnostic and outcome assessment protocols (see ***Table 2***). This will allow, for example, genomic analyses based on particular subgroups (e.g., tic-related OCD, familial vs sporadic, etc.) or the use of polygenic scores to predict disease severity.

**Table 2.**
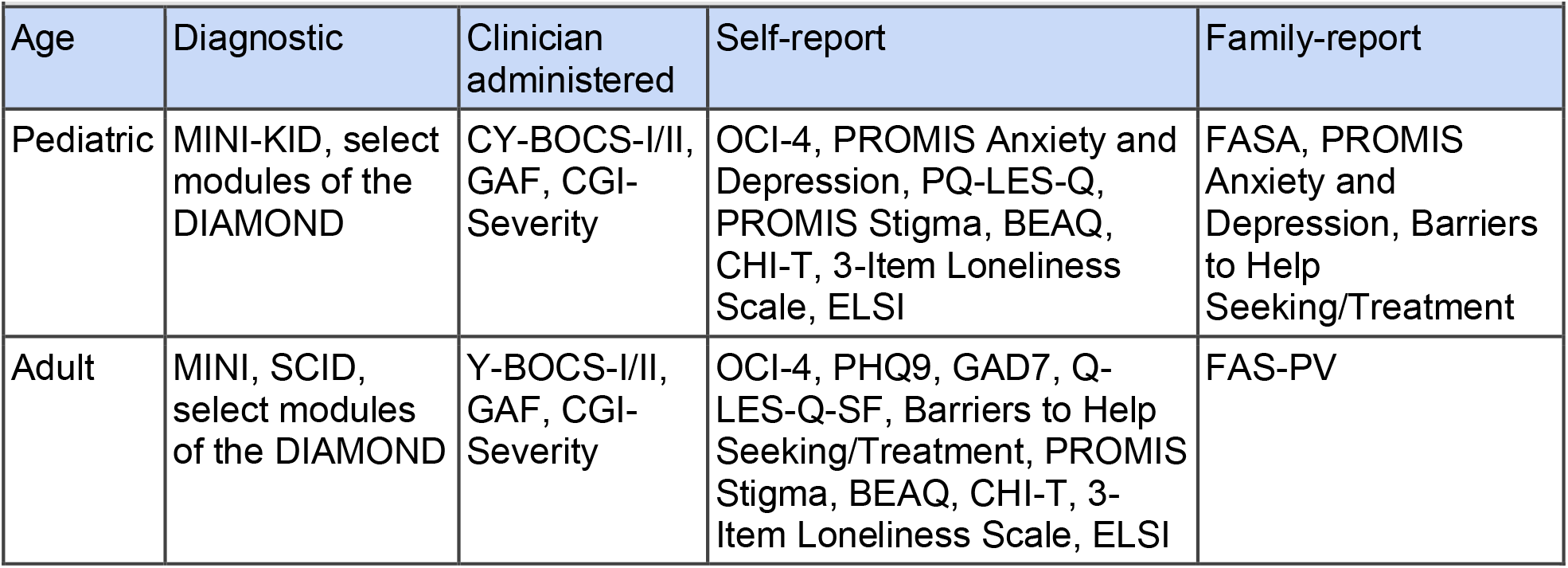
Harmonized core assessment measures used across all collection sites

|  |  |  |  |  |
| --- | --- | --- | --- | --- |
| Age | Diagnostic | Clinician administered | Self-report | Family-report |
| Pediatric | MINI-KID, select modules of the DIAMOND | CY-BOCS-I/II, GAF, CGI-Severity | OCI-4, PROMIS Anxiety and Depression, PQ-LES-Q, PROMIS Stigma, BEAQ, CHI-T, 3-Item Loneliness Scale, ELSI | FASA, PROMIS Anxiety and Depression, Barriers to Help Seeking/Treatment |
| Adult | MINI, SCID, select modules of the DIAMOND | Y-BOCS-I/II, GAF, CGI-Severity | OCI-4, PHQ9, GAD7, Q-LES-Q-SF, Barriers to Help Seeking/Treatment, PROMIS Stigma, BEAQ, CHI-T, 3-Item Loneliness Scale, ELSI | FAS-PV |

All assessments were subject to a thorough translation, back translation, and cross-cultural adaptation and validation process in order to guarantee construct validity. All instruments were available in English. Bibliographic research was carried out in order to investigate the availability of each instrument in Spanish and Portuguese, and whether there were relevant reliability and validity studies.

Instruments unavailable in the languages needed for the study (Spanish or Brazilian Portuguese) were selected for a cross-cultural validation process, in order to guarantee the instrument’s construct validity. Parameters applied were derived from the Consensus-based Standards for the Selection of Health Measurement Instruments (COSMIN) study design checklist for patient-reported outcome measurement instruments (Mokkink et al., 2019; Terwee et al., 2018). First, independent translations were performed by two investigators. Second, a consensus was made by one specialist and discordances were reviewed by a second specialist. Third, a back-translation was performed by an English-certified proficient investigator. Fourth, the back translation was evaluated by the author of the original instrument and after the author’s suggestions and approval, adjustments were made to develop the final version. Fifth, the final version of the instrument was submitted to the test phase where ~40 participants were interviewed, even for self-report measures, about the clarity and accuracy of the instruments. Final instrument adjustments were made according to the test phase results.

The core battery includes a variety of clinician-administered, self-report, and parent-report (for children) measures. The Mini International Neuropsychiatric Interview (MINI)(Sheehan et al., 1998), the Structured Clinical Interview for DSM-V (SCID)(First & Williams, 2016), and MINI-KID(Sheehan et al., 2010) are focused, structured diagnostic interviews that assess DSM-5 and ICD-10 psychiatric disorders in adults and youth. The Children’s Yale-Brown Obsessive Compulsive Scale (CY-BOCS-I/II)(Scahill et al., 1997; Storch et al., 2019) and the Y-BOCS-I/II(Goodman et al., 1989; Storch et al., 2010) are clinician-rated, semi-structured interviews that assess presence and severity of OCD symptoms in youth and adults. The Obsessive-Compulsive Inventory-4 (OCI-4)(Abramovitch et al., 2021) is a self-report measure that assesses obsessive-compulsive symptom dimensions in youth and adults. Modules of the Diagnostic Interview for Anxiety, Mood, and OCD and Related Neuropsychiatric Disorders (DIAMOND)(Tolin et al., 2018) are used to assess the presence of obsessive-compulsive related disorders (OCRD). The Global Assessment of Functioning (GAF)(Schwartz & Del Prete-Brown, 2003) scale is a single, clinician-rated item assessing the overall severity of psychopathology and functional impairment in the worst week of the past month. The Clinical Global Impression-Severity (CGI-S)(Guy, 1976) is a clinician-rated assessment of clinical severity.

Self-reported depressive and anxiety symptoms are assessed with the PHQ-9(Kroenke et al., 2001) and GAD-7(Spitzer et al., 2006) for adults and the PROMIS Depression and Anxiety scales (youth)(Quinn et al., 2014). PROMIS Stigma(Molina et al., 2013) measures experiences with stigma related to OCD in both adults and youth. The Family Accommodation Scale - Patient Report (adults)(Wu et al., 2016) and Family Accommodation Scale - Anxiety (youth)(Lebowitz et al., 2013, 2015) are used to assess family accommodation to the OCD symptoms. The Quality of Life Enjoyment and Satisfaction Questionnaire - Short Form (Q-LES-Q-SF)(Endicott et al., 1993) and PQ-LES-Q(Endicott et al., 2006) are self-report measures that assess satisfaction across physical health, mental health, and well-being in adults and youth. The Barriers to Help Seeking/Treatment(Goodwin et al., 2002; Marques et al., 2010) examines factors that prevent one from seeking treatment. The Brief Experiential Avoidance Questionnaire (BEAQ)(Gámez et al., 2014) is included to assess experiential avoidance in both adults and youth. To measure self-reported compulsivity and associated functional impairment, the Cambridge-Chicago Compulsivity Trait Scale (CHI-T)(Chamberlain & Grant, 2018) is used. The Three-Item Loneliness Scale(Hughes et al., 2004) is a self-report measure that captures feelings of loneliness. To address ethical, legal, and social aspects of this work, we developed a short questionnaire that measures participants’ attitudes toward genetic testing for psychiatric conditions. Beyond these core assessments, a supplemental battery of measures has been collaboratively generated. These measures are administered on a rotational basis throughout the study to minimize burden while capturing important clinical constructs.

Each participant is comprehensively assessed in person or via a secure video conferencing system in the language the person feels most comfortable using. All symptom-oriented assessments are conducted by trained raters at either the recruitment site or by Baylor College of Medicine (BCM) personnel. Cross-site training includes a review of each measure and co-rating videotaped cases. For example, raters view several videotaped ratings of the (C)Y-BOCS-I/II and then independently rate several tapes in their respective language. Raters must score within 20% of the gold standard on the (C)Y-BOCS-I/II. Regular BCM-led office hours are held to complement on-site supervision. A (C)Y-BOCS-I/II expert committee comprised of a subset of site PIs was formed in order to review and score all (C)Y-BOCS-I/II assessment training videos to ensure uniformity in administration and scoring. These expert committee members also made themselves available for (C)Y-BOCS-I/I consultation with other PIs and assessors.

#### 2.1.6 Demographic assessments

In addition to collecting core demographic information (e.g., age, sex, education, family composition, etc.), comprehensive data about past diagnoses and treatment history are collected. Psychotherapy history, particularly a course of exposure and response prevention, is assessed, as well as the associated treatment response. Detailed information regarding medication history is collected, including past medication trials, highest dose, duration of treatment, side effects, and degree of symptom improvement. Barriers to treatment, social impact, and perceptions of reproductive decision-making are also collected.

#### 2.1.7 Clinical data collection and quality assurance

Phenotypic data is collected via the HIPPA-compliant Research Electronic Data Capture (REDCap), a secure web-based survey collection, direct entry platform, and database (Harris et al., 2009, 2019). Data quality is assessed by including REDCap flags and manual data checks. Automated data checks are planned using an R-Studio API with REDCap. Cross-site data will be compared for missing data and inter-question correlation to determine consistency. Study assessments are audio recorded and reviewed for interrater reliability. Site PIs provide (C)YBOCS-I/II and MINI-KID/MINI supervision in order to ensure accurate assessment administration and review recorded audio of (C)YBOCS-II and MINI-KID/MINI administration, as needed. When assessment and scoring remain unresolved during local site supervision, assessors will bring concerns to the expert committee to help come to a consensus about assessment scoring.

### 2.2 Aim 2

#### 2.2.1 Genotyping

At the time of this writing, our plan is to genotype all 5,000 Latinx OCD cases with the Illumina Global Screening Array (GSA), which has been shown to adequately capture variation >1% across all populations in the Thousand Genomes Project, and match with data from at least 10,000 Latinx controls planned to be genotyped on the same array. If, however, costs substantially decrease for a more comprehensive methodology (e.g., low-pass whole genome plus exome sequencing or standard whole genome sequencing), we will seek to change our plan. This is particularly true if the controls on which we rely are interrogated with a different method. For the purposes of this study protocol, we focus on a scenario involving GSA genotyping, given that this is the current plan.

#### 2.2.2 Admixture and *Tractor*

“Admixed” individuals possess DNA with more than one ancestral component. Admixed individuals, including those from Latin America, have been systematically excluded from genetic studies due to the historic lack of available tools to appropriately handle their complex genomes. If not accounted for, such population substructure can compromise analyses and bias results when highly admixed samples are included alongside more homogenous individuals using standard analysis (Sul et al., 2018). In GWAS, the specific concern regarding including admixed participants is obtaining false positive associations due to alleles being at different frequencies across populations. Previous studies have attempted to control for this by using principal components (PCs) in linear or linear mixed model frameworks. However, PCs capture only broad admixture fractions, and individuals’ local ancestry makeup may differ between case and control cohorts even if their global fractions are identical. Including PCs as covariates using standard GWAS methods still leaves open the possibility for false positive associations in admixed cohorts.

To conduct gene discovery in the best-calibrated manner for these Latinx cohorts, we will utilize the *Tractor* framework (Atkinson et al., 2021). *Tractor* corrects for fine-scale population structure at the level of haplotypes by using local ancestry, allowing admixed samples of all mixture proportions to be readily included in large-scale collections in a well-calibrated manner. *Tractor* also has been shown to boost GWAS power in admixed cohorts to identify ancestry-specific signals. Furthermore, *Tractor* produces ancestry-specific effect size estimates and p values, which other strategies such as classic GWAS or admixture mapping can not. These ancestry-specific effect sizes will be extremely valuable to downstream efforts, such as in fine-mapping and building genetic risk scores (see below).

#### 2.2.3 LATINO GWAS

First, we will perform extensive quality control (QC) of the genotyping data as described in previous work from the PGC (Howard et al., 2019; Trubetskoy et al., 2022). Second, we will match cases and controls through an iterative procedure meant to minimize the chances of population stratification. Third, we will quantify the ancestral composition of each subject in order to appropriately select reference panels for local ancestry inference. Fourth, we will perform imputation using reference sets from the Trans-Omics for Precision Medicine (TOPMed) program, which has whole genome sequencing data from a large, diverse set of samples. Fifth, we will identify ancestral tracts by performing local ancestry inference using RFmix_v2 (Maples et al., 2013).

These five steps will produce the input data needed to perform a case-control GWAS using *Tractor*. We will include standard covariates plus an estimate of global ancestry to control for any phenotypic effect due to overall admixture makeup. The results will be combined using a model that doesn’t assume the same effect size across ancestries. Several methods have been designed for cross-ancestry meta-analysis that allow for effect size heterogeneity (e.g., MANTRA, MR-MEGA, MAMA). For chrX, we will use an additive logistic regression model with the same covariates. We will test the genome-wide distribution of the test statistic in comparison with the expected null distribution using the genomic inflation factor λGC and QQ plots. λGC quantifies the extent of the bulk inflation resulting from a combination of true polygenic signal, systematic technical bias and population stratification (Devlin & Roeder, 1999). In order to quantify the contribution of these factors, we will use LD Score regression (Bulik-Sullivan et al., 2015), where the intercept estimates the inflation in the mean chi-square that results from confounding biases, such as cryptic relatedness or population stratification. To declare genome-wide significance, we will strictly adhere to the field-standard P-value threshold of 5×10^−8^ (Pe’er et al., 2008).

A major strength of this study is the depth and consistency of phenotyping across sites (***Table 2***), allowing us to analyze the genetics of biologically plausible subtypes (early onset, tic-related) and symptom dimensions. As opposed to the case-control GWAS, these analyses will consider both quantitative, as well as categorical data, and are restricted to cases only.

#### 2.2.4 Trans-ancestry GWAS with PGC OCD

Most standard GWAS meta-analysis methods assume fixed-effects models that predict variants to have the same true marginal effect size across all studies. This is not necessarily a proper assumption in trans-ancestry meta-analyses. Although studies to date have generally shown that the causal genetic effect of a variant is constant across populations, there are multiple reasons for differences in effect size estimates across populations. For example, different LD structures across populations, differences in gene-gene interactions or gene-environment interactions, and phenotypic differences may produce heterogeneity across cohorts. Therefore, it is advised to model this cross-cohort heterogeneity in meta-analysis by using a random effects model. This statistical approach allows for the true marginal effect size of each variant to vary across studies, testing the null hypothesis that the mean true effect size across cohorts is zero. Alternatively, a transancestral meta-analysis model that assumes variant effect sizes may differ more between more genetically diverse populations can be used. Statistical methods appropriate to test our study aims are actively in development and we will ensure that our meta-analytic approach for PGC OCD is appropriate.

LATINO will triple the current number of non-EA OCD cases available for GWAS with PGC OCD. Currently, PGC OCD has >50,000 cases, only ~2,500 of which are of non-EA. We anticipate that over the next five years the PGC sample will grow to around 80,000 cases, with contributions from ongoing clinical collections and large biobanking efforts such as the Million Veteran Program (MVP)(Gaziano et al., 2016) and the AllofUs Research Program (All of Us Research Program Investigators et al., 2019). These latter two efforts in particular should help to add more non-EA OCD cases to the pool available to PGC OCD, but there will still be a heavy bias toward EA individuals. Therefore, this study will help to ensure that non-EA populations are not left behind as OCD genomics advances.

### 2.3 Aim 3

#### 2.3.1 Fine-mapping

Genome-wide significant loci are typically large in size (tens of kilobases to over a megabase) and include many variants with similar *p* values. The causal variant(s) within GWAS peaks are often difficult to separate from those that are in LD. Fine-mapping is used to decrease the search space for the causal variant(s) and facilitate follow-up or functional genomic work. Recent work(Lam et al., 2019; Ng et al., 2017; Wyss et al., 2018) has shown that genetic diversity can be used to improve fine-mapping resolution, owing to differences in the pattern of LD between causal and non-causal variants. If we assume that causal variants will have consistent effects across populations and non-causal variants will have inconsistent effects due to population-specific LD, then we would expect causal variants to be more statistically significant and less heterogeneous in a trans-ancestry meta-analysis compared to non-causal variants (Peterson et al., 2019). In addition, haplotype block size may be smaller in the non-European ancestry groups, improving resolution.

The development of new methods for fine-mapping, particularly trans-ancestry fine-mapping, is a very active field of research at present. Different methodologies have particular strengths and weaknesses. We will utilize SuSiEx (SUm of SIngle Effects trans-ancestry), which is a new cross-population fine-mapping extension of SuSiE (Zou et al., 2022). We will fine-map any locus that reaches genome-wide significance in the transancestry meta-analysis or in ancestry-specific analyses. We will pull out a 100 kb contiguous region centered on the most significant SNP (50 kb on either side). This is a conservative choice, as LD generally begins to decay between SNPs separated by >25 kb (Conrad et al., 2006). We will integrate functional annotation data, since multiple studies have shown this can improve statistical fine-mapping (Kichaev et al., 2014; Kichaev & Pasaniuc, 2015). There also exists substantial functional annotation data for brain tissue and brain cells(Wang et al., 2018) that we can use for this project. Many locus-specific properties will influence our ability to fine-map a particular locus at high resolution. These include the number of causal SNPs and differences between populations in minor allele frequency, LD structure and SNP density (Schaid et al., 2018).

#### 2.3.2 Trans-ancestry polygenic risk score analysis

Polygenic risk scores (PRS) are used to predict complex traits using genetic data. Most PRS methods are inadequate in non-EA populations because of decreased prediction accuracy, possibly due to different patterns of LD, allele frequencies, or overall genetic architecture (Martin et al., 2020). This limits their clinical use for precision medicine efforts in non-European populations. As with fine-mapping, the development of new methods for trans-ancestry PRS work is a very active field of research.

A new PRS construction method, PRS-CSx, has recently been shown to improve cross-population polygenic prediction by integrating GWAS summary statistics from multiple populations. This allows for the benefits of including the largest discovery GWAS (typically on Europeans), as well as a better-matched discovery sample of more modest size. PRS-CSx(Ruan et al., 2022) couples genetic effects across populations via a shared continuous shrinkage (CS) prior, enabling more accurate effect size estimation by sharing information between summary statistics and leveraging linkage disequilibrium diversity across discovery samples. PRS-CSx has shown improved prediction performance across traits with a wide range of genetic architectures, cross-population genetic overlaps and discovery GWAS sample sizes.

Recent multi-ancestral PRS studies have demonstrated that working with limited sample sizes for non-European cohorts is still valuable, as it allows for larger than expected gains in predicting polygenic traits (Bigdeli et al., 2020; Lam et al., 2019). We anticipate that further increases in the non-European sample size will lead to ever larger gains in variance explained and, ultimately, to the clinical utility of PRS across populations.

## 3 DISCUSSION

While new discoveries are taking place in the genetic understanding of OCD, most of these advances are built upon exclusively EA samples. For example, we are very encouraged by the recent discovery of the first 30 genome-wide significant hits for OCD, but this was achieved with an entirely EA sample, limiting the utility of these results in non-EA cases and exacerbating health disparities. The LATINO study provides the opportunity to meaningfully address this issue, which has critical implications for understanding of disease pathophysiology (e.g., identifying causal variants), and ensuring accurate PRS modeling. Beyond the contribution to OCD genetics research, this collaborative study may serve as a model for engaging stakeholders in research.

There is a pressing need for more accessible evidence-based treatments for OCD throughout the world, including Latin America. Therefore, one of the major goals of LATINO is capacity building. In collaboration with the International OCD Foundation, we have already established clinical consultation groups and a Spanish-language exposure and response prevention training institute. Furthermore, the Asociación Latinoamericana de Trastorno Obsesivo Compulsivo (ALTOC) and LATINO recently hosted the very first OCD conference ever held in Latin America. This event occurred in June 2022 in Cartagena, Colombia under the name “LATINO Cartagena: The 1st Latin American Congress on OCD”. This meeting was a great success with >250 attendees (face-to-face and virtual), with >90% of attendees being of Latin American ancestry. One of the unique aspects of the meeting was its broad audience - a mixture of researchers, mental health professionals, students, and people with OCD and their families. Two tracks were available to participants (one for patients and families and a second for students/researchers and clinicians), which ensured that the congress had something to offer to all attendees.

We have now established this as an annual educational meeting that will rotate throughout Latin American countries. Given the significant disability associated with OCD, increasing the number of trained clinicians and thereby improving access to evidence-based care has the potential to reduce suffering for many individuals worldwide. Furthermore, developing academic opportunities for publications, grant applications, and presentations is a core focus of this project. Mentoring relationships have and will continue to be established with experienced investigators throughout the consortium with the goal of developing the next generation of scholars and clinicians.

LATINO has a number of future directions. First, although our assessment battery is uniquely comprehensive in assessing obsessive-compulsive and related symptoms, a number of constructs were not included. We are hoping for the opportunity to integrate measures of cognition, anhedonia, and suicidality. Second, we hope to generate more comprehensive genomic data. We are funded to perform microarray-based genotyping but are seeking opportunities to whole exome or whole genome sequence LATINO samples. Third, we are primarily collecting samples from individual OCD cases, but are pursuing opportunities to also collect samples and data from biological parents and other family members to facilitate trio and other family-based genetic study designs. Finally, LATINO welcomes further collaboration with anyone interested in OCD in Latinx individuals and is proud to provide an infrastructure to explore new research ideas to help this underserved population.

## Data Availability

Our liberal data and analysis sharing principles will make phenotypic and genotype data and scripts widely available for access by other scientists to maximize the utility of our investigation. The datasets generated and analyzed will be available in the NIMH Data Archive (https://nda.nih.gov/). Genomic data will also be available through the Psychiatric Genomics Consortium (PGC, https://pgc.unc.edu/). When permitted by the local IRB, DNA samples will be available from the NIMH Repository and Genomics Resource (https://www.nimhgenetics.org/).

## ACKNOWLEDGMENTS

The LATINO team acknowledges the contributions of Dave Keller, Esme Rosario, Natasha Sefcovic, Emily O’Bryant, Katia Regina da Silva, Cynthia Bulik, Stephanie Crowley, Leanne Scott, Brandy Duke, Katy Gathron, Duff Dean, McKenzie Sluder and Cole Whiteman for their input on various aspects of the project. We would like to acknowledge Gracielle Nonato for help translating instruments into Portuguese and members of the University of Puerto Rico Center for the Study and Treatment of Fear and Anxiety for their work on this project. Study data are collected and managed using REDCap electronic data capture tools provided by the Baylor College of Medicine (BCM). We’d also like to thank Uma Ramamurthy and her team at BCM Research IT for designing the LATINO study website. All LATINO sites thank, in particular, the essential contribution of those with lived experience and their families for participating in LATINO.

## ETHICS APPROVALS

LATINO was approved by the Baylor College of Medicine (BCM) Institutional Review Board (IRB) (Approval Number: H-49814) under the single IRB (sIRB) framework. BCM is the IRB of record for all US-based sites. Each site outside the US has received ethical approval from a local IRB.

## CONSORTIA

### Members of the Latin American Trans-ancestry INitiative for OCD genomics (LATINO)

Jonathan S Abramowitz, Victor R Adorno, Cinthia Aguirre, Cynthia Alvarado, Gustavo S Alves, Gilberto S Alves, NaEshia Ancalade, Jacey L Anderberg, Alejandro A Arellano Espinosa, Paul D Arnold, Elizabeth G Atkinson, Daphne M Ayton, Izabela G Barbosa, Laura Marcela Barón Castano, Cynthia N Barrera, Liz Basanez, María Celeste Berardo, Dayan Berrones, John R Best, Tim B Bigdeli, Tania Borda, Christie L Burton, Jennifer L Callahan, Carolina Cappi, Maria Cecília B Carneiro, Sandra L Cepeda, Evelyn Chazelle, Jessica Marianela Chire Alvarez, Macarena Churruca Munoz, Pamela Claisse Quiroz, Journa Cobite, Jonathan S Comer, Daniel L Costa, Jennifer Crosbie, James J Crowley, Victor O Cruz, Guillermo Dager, Luisa F Daza, Anabel De la Rosa, Daniela del Río Zamacona, Fernanda Z Delage, Carolina B Dreher, Lucila Fay, Tomas Fazio, Ygor A Ferrão, Gabriela M Ferreira, Edith Figueroa, Leonardo F Fontenelle, Diego A Forero, Daniele TH Fragoso, Renee M Frederick, Bharathi S Gadad, Sheldon R Garrison, Paola Giusti-Rodriguez, Laura D Gonzalez, Marco A González, Polaris Gonzalez-Barrios, Wayne K. Goodman, Daniel G Guttfreund, Andrew G Guzick, Matthew W Halvorsen, Joseph D Hovey, Reinhard Janssen-Aguilar, Matias Jensen, Joali Juárez Lujambio, Nasim Khalfe, Madison A Knutsen, Minjee Kook, Nuria Lanzagorta, Gabriel Lázaro-Muñoz, Monicke O Lima, Melanie O Longhurst, Jessica Lopez-Harder, David A Lozada Martinez, Elba Susana Luna, Andrea H Marques, Molly Martinez, Mayra C Martinez Mallen, Karen G Martinez-Gonzalez, Maria de Los Angeles A Matos, Manuel Mattheisen, Caitlyn E Maye, Joseph F McGuire, Gabriela B de Menezes, Euripedes C Miguel, Charlene Minaya, Tomás Miño, Sara M Mithani, Circe Montes de Oca, Alonso Morales-Rivero, Maria E Moreira-de-Oliveira, Olivia J Morris, Pablo R Moya, Maria Beatriz Moyano, Sandra I Muñoz, Zainab Naqqash, Humberto Nicolini, Gracielle C Nonato, AmbarA Nunez Bracho, Belinda E Nunez Bracho, Maria Corina Ochoa Rojas, Marcos E Ochoa-Panaifo, Luis A Olavarria Castaman, Iliana Ortega, Darpan I Patel, Michele T Pato, Ainsley K Patrick, Mariel Paz y Mino, Jose L Perales Orellana, Tamara Peregrina, Stacey Pereira, Tania Pérez Duarte, Kelly L. Piacsek, Maria Belen Prieto, Lucas de C Quarantini, Yana Quarantini-Alvim, Renato T Ramos, Iaroslava C Ramos, Vanessa R Ramos, Kesley A Ramsey, Diana Rancourt, Elise V Ray, Alexandra Z Jimenez-Reynolds, Margaret A Richter, Bradley C Riemann, Juan C Rivas, Carolyn I Rodriguez, Maria C Rosario, Angel A Ruiz-Chow, Alejandra Ruiz-Velasco, Aline S Sampaio, Leonardo C Saraiva, Russell J Schachar, Sophie C Schneider, Ethan J Schweissing, Laura D Seligman, Roseli G Shavitt, Keaton J Soileau, S Evelyn Stewart, Eric A Storch, Shaina B Storch, Emily R Strouphauer, Kiara R Timpano, Beatriz Treviño de la Garza, Javier Vargas-Medrano, Guadalupe Vidal Martinez, Saira A Weinzimmer, Andrew D Wiese, Mauricio A Yanez, Gwyneth Zai, Lina M Zapata-Restrepo, Luz M Zappa, Raquel M Zepeda-Burgos, Anthony W Zoghbi

### Members of the Brazilian Obsessive-Compulsive Spectrum Disorder Working Group (GTTOC)

Aline Santos Sampaio, Ana Gabriela Hounie, Ana Paula Ribeiro, Angela Rodrigues Souza e Silva, Ariane Fadel Martinho, Bárbara Luciane Perdigão Stumpf, Bianca Torres Mendonça de Melo Fadel, Camila Vale Porto, Carina Freitas Chacur, Carla Pereira Loureiro, Carolina Blaya Dreher, Carolina Cappi,, Cristiane Flôres Bortoncello, Daniel Lucas da Conceição Costa, Daniele Tamae Hashimoto Fragoso, Euripedes Constantino Miguel, Fernanda Zetola Delage, Flávia Cervone, Gabriela Bezerra de Menezes, Gabriela Cirqueira de Souza Barros, Gabriela Mourão Ferreira, Gilberto Sousa Alves, Gustavo Santiago Alves, Hugo Caminha, Izabela Guimarães Barbosa, Jaqueline Cenci, Julia Fernandes Eigenheer Muhlbauer, Juliana Rigue da Silva, Leonardo Cardoso Saraiva, Leonardo Franklin da Costa Fontenelle, Lígia S. Camba Lopez, Livi Ferreira Testoni de Faro, Luan Pinheiro Domingues Moreira, Luana Dumans Laurito, Lucas de Castro Quarantini, Luciana Midori Samezima, Luiz Felipe Guimarães dos Santos Martins, Marcos Vinícius Sousa de Oliveira, Maria Alice de Mathis, Maria Carolina Paraíso Lopes, Maria Cecília Carneiro, Maria Conceição do Rosário, Maria Eduarda Ismerio Moreira de Oliveira, Marina Martorelli Pinho, Monicke de Oliveira Lima, Nádia Nagano, Patrycia Chedid Danna, Pedro Pereira Fortes da Silva, Raquel Fernandes de Jesus Cardoso, Renan Barbosa de Christo, Richard Chuquel Silveira de Avila, Rodrigo Bolsson Radins, Roseli Gedanke Shavitt, Samara dos Santos Ribeiro, Tatiane Veríssimo da Silveira Meirelles, Thays Mendes Aguiar, Theresa Kerolayne da Silva Sousa, Thiago Blanco Vieira, Vanessa Rogério Ramos, Veronica Huhne de Vasconcellos, Yana Quarantini Alvim, Ygor Arzeno Ferrão.

## ON-LINE RESOURCES

LATINO study website: www.latinostudy.org

